# Is using Dual Energy Virtual Enhancing over Non-Contrast CT beneficial in terms of Diagnostic Accuracy of finding Adrenal Adenoma? : A Meta-analysis

**DOI:** 10.1101/2023.07.13.23292609

**Authors:** Dev Desai, Arman Duisenbayev, Pankti Maniyar, Maria Eleni Malafi, Dev Andharia, Abhijay B. Shah

**Author notes:** (Corresponding author) Email Address, Mob No.

## Abstract

**Background:** The study compared the diagnostic accuracy of Dual Energy Virtual Non-Contrast CT (DEvNCT) to Non-Contrast CT (NCT) for adrenal adenomas. Adrenal nodules on CT scans require precise characterization to differentiate between benign and malignant lesions. CT imaging uses features like low attenuation and contrast enhancement to identify adenomas. However, distinguishing adenomas from non-adenoma lesions can be challenging due to overlapping attenuation levels.

DEvNCT, a post-processing technique enabled by dual-energy CT, optimizes adrenal lesion evaluation. The study aimed to compare the diagnostic accuracy of DEvNCT and NCT for adrenal adenomas.

**Methodology:** A systematic review and meta-analysis searched relevant literature in PubMed, Google Scholar, and Cochrane Library databases. The analysis included nine randomized controlled trials (RCTs) with 407 patients: three RCTs for NCT vs. Biopsy and six RCTs for DEvNCT vs. NCT. Study quality was assessed using appropriate tools.

**Result:** Results revealed DEvNCT had 72% sensitivity, 94% specificity, and 0.878 positive predictive value for adrenal adenoma diagnosis. NCT had 80% sensitivity, 68.1% specificity, and 0.831 positive predictive values. The difference in specificity between DEvNCT and NCT was significant, indicating DEvNCT’s superior ability to identify adenomas accurately.

**Conclusion:** The study concluded that DEvNCT exhibited comparable sensitivity but higher specificity than NCT in diagnosing adrenal adenomas. DEvNCT could potentially reduce follow-up imaging, lower costs, and minimize radiation exposure. However, biopsy remains the gold standard for confirming adrenal adenoma diagnosis.

Overall, DEvNCT shows promise for improving diagnostic accuracy compared to NCT in diagnosing adrenal adenomas. Further research and clinical validation are needed to confirm these findings.

## Introduction

Adrenal nodules are one of the frequent incidental imaging findings, which are identified in approximately 5% of patients undergoing computed tomography (CT). [1], [2]. The majority of adrenal lesions discovered incidentally are benign adenomas. However, the challenge lies in distinguishing between adenomas that are benign and malignant, especially in patients with a history of malignancy outside the adrenal glands. Therefore, it is crucial to accurately characterize incidentally detected adrenal nodules through imaging [3]– [5].

Low attenuation on unenhanced scans due to the presence of intracytoplasmic fat, and maximal contrast enhancement during the portal venous phase followed by rapid washout during the delayed phase are two important CT imaging features of adrenal adenomas.[6]– [14]. The presence of intracytoplasmic lipids in adrenal cortical cells is a crucial factor in steroid production and can be found in approximately 70% of adenomas[7]. The identification of intracytoplasmic lipids in an adrenal nodule through CT or MRI imaging is the foundation for diagnosing “lipid-rich” adenomas. Conversely, “lipid-poor” adenomas can be identified by their high relative washout rate during a CT study with a delayed contrast phase [15]. Intracytoplasmic lipids with a threshold of 10 Hounsfield units on non-contrast CT can confirm the presence of adrenal adenoma with high sensitivity and specificity. For adrenal lesions < 4 cm and with density < 10 HU, further imaging is not required. However, detecting incidental adrenal nodules during a single-phase contrast-enhanced CT can lead to a common clinical dilemma due to the overlap in enhanced adrenal nodule attenuation levels between adenomas and non-adenoma lesions [16]– [19]. Additional diagnostic tests, such as dedicated multiphasic CT, chemical shift MRI, fluorine 18 (18F) fluorodeoxyglucose (FDG) PET/CT, interval follow-up, or percutaneous biopsy in conjunction with laboratory tests, may be necessary in such cases. However, most of the lesions eventually turn out to be benign adenomas [6], [20].

Several postprocessing techniques enabled by Dual-energy virtual non-contrast CT (DEvNCT) can provide an optimized evaluation of adrenal lesions. Unenhanced reconstructions have shown comparable sensitivity to true unenhanced images in distinguishing between benign and malignant adrenal nodules [20]. Iodine quantification using dual-energy CT can be used as an alternative way to measure tissue perfusion. Using virtual non-contrast CT (vNCT) to diagnose adenomas is desirable as it could eliminate the need for further imaging. However, studies reporting the accuracy of vNCT for diagnosis using absolute attenuation are limited. This systematic review aims to compare the diagnostic accuracy of virtual non-contrast CT (vNCT) to non-contrast CT (NCT) in the diagnosis of adrenal adenomas.

## METHODOLOGY

### DATA COLLECTION

For the collection of the data, a search was done by two individuals using PubMed, Google Scholar, and Cochrane Library databases for all relevant literature. Full - Text Articles written only in English were considered.

The medical subject headings (MeSH) and keywords ‘Dual-Energy CT(DECT) For Adrenal Adenoma’, ‘Non-contrast CT (NCT\NCCT) for Adrenal Adenoma’, ‘Biopsy of Adrenal gland lesions’, and ‘Adrenal gland imaging for Adenomas’ were used. References, reviews, and meta-analyses were scanned for additional articles.

### INCLUSION AND EXCLUSION CRITERIA

Titles and abstracts were screened, and Duplicates and citations were removed. References of relevant papers were reviewed for possible additional articles. Papers with detailed patient information and statically supported results were selected.

We searched for papers that show more accurate diagnoses, where procedures considered were DECT and NCT.

The inclusion criteria were as follows: (1) studies that provided information about the accurate diagnosis with DECT and NCT; (2) studies published in English; (3) Studies comparing DECT with NCT and NCT with Biopsy as a Diagnosis modality for Adrenal gland Adenomas.

The exclusion criteria were: (1) articles that were not full text, (2) unpublished articles, and (3) articles in other languages.

### DATA EXTRACTION

Each qualifying paper was independently evaluated by two reviewers. Each article was analyzed for the number of patients, their age, procedure modality, and incidence of the predecided complications. Further discussion or consultation with the author and a third party was used to resolve conflicts. The study’s quality was assessed using the modified Jadad score. In the end, According to PRISMA, a total of 8 RCTs with a total of 407 patients were selected, out of which 3 RCTs with a total of 139 patients were selected for NCT vs Biopsy study and 6 RCTs with a total of 268 patients were selected for DECT vs NCT study. Ho et al. 2012 were common for both analyses.

### ASSESSMENT OF STUDY QUALITY

Using the QualSyst tool, two writers, independently assessed the caliber of each included study. This test consists of 10 questions, each with a score between 0 and 2, with 20 being the maximum possible overall score. Two authors rated each article independently based on the above criteria. The interobserver agreement for study selection was determined using the weighted Cohen’s kappa (K) coefficient. For deciding the bias risk for RCTs, we also employed the Cochrane tool. No assumptions were made about any missing or unclear information. there was no funding involved in collecting or reviewing data.

### STATISTICAL ANALYSIS

The statistical software packages RevMan (Review Manager, version 5.3), SPSS (Statistical Package for the Social Sciences, version 20), Google Sheets, and Excel in Stata 14 were used to perform the statistical analyses. The data was obtained and entered into analytic software [21]. Fixed- or random-effects models were used to estimate Sensitivity, Specificity, positive predictive value (PPV), diagnostic odds ratios (DOR), and relative risk (RR) with 95 percent confidence intervals to examine critical clinical outcomes (CIs). Diagnosis accuracy and younden index were calculated for each result. Individual study sensitivity and specificity were plotted on Forest plots and in the receiver operating characteristic (ROC) curve. The forest plot and Fagan’s Nomogram were used to illustrate the sensitivity and specificity of different papers.

### BIAS STUDY

The risk of bias was evaluated by using QUADAS-2 analysis. This tool includes 4 domains as Patient selection, Index test, Reference standard, Flow of the patients, and Timing of the Index tests.

## RESULT

### DECT v NCT

Here, Table 1 describes all the description of papers used for the DECT vs NCT study. As the result described above, in the forest chart (figure 2), the comparison of the sensitivity and specificity of different papers can be seen. The same is illustrated in the SROC curve(figure3). A total of 6 RCTs with 268 patients were selected for the study. Out of which 1 study showed sensitivity above 95%. And 4 studies showed Specificity above 95%. And 1 study showed sensitivity and specificity both above 95%. The value of True positive was 72, True Negative was 158, False negative was 28, and False Positive was 10. With a confidence interval of 95%, Sensitivity, specificity, and Positive Predictive values were calculated. A summary of this is available in Figure 2. The Sensitivity of the DECT is 0.72 with a CI of 95% in a range of (0.553 to 0.887) the mean being (0.167). The Specificity of the DECT is 0.94 with a CI of 95% in a range of (0.871 to 1.01) the mean being (0.214). The positive predictive value (PPV) is 0.878 with a CI of 95% in a range of (0.664 to 1.092) the mean being (0.214).

**Table 1:**
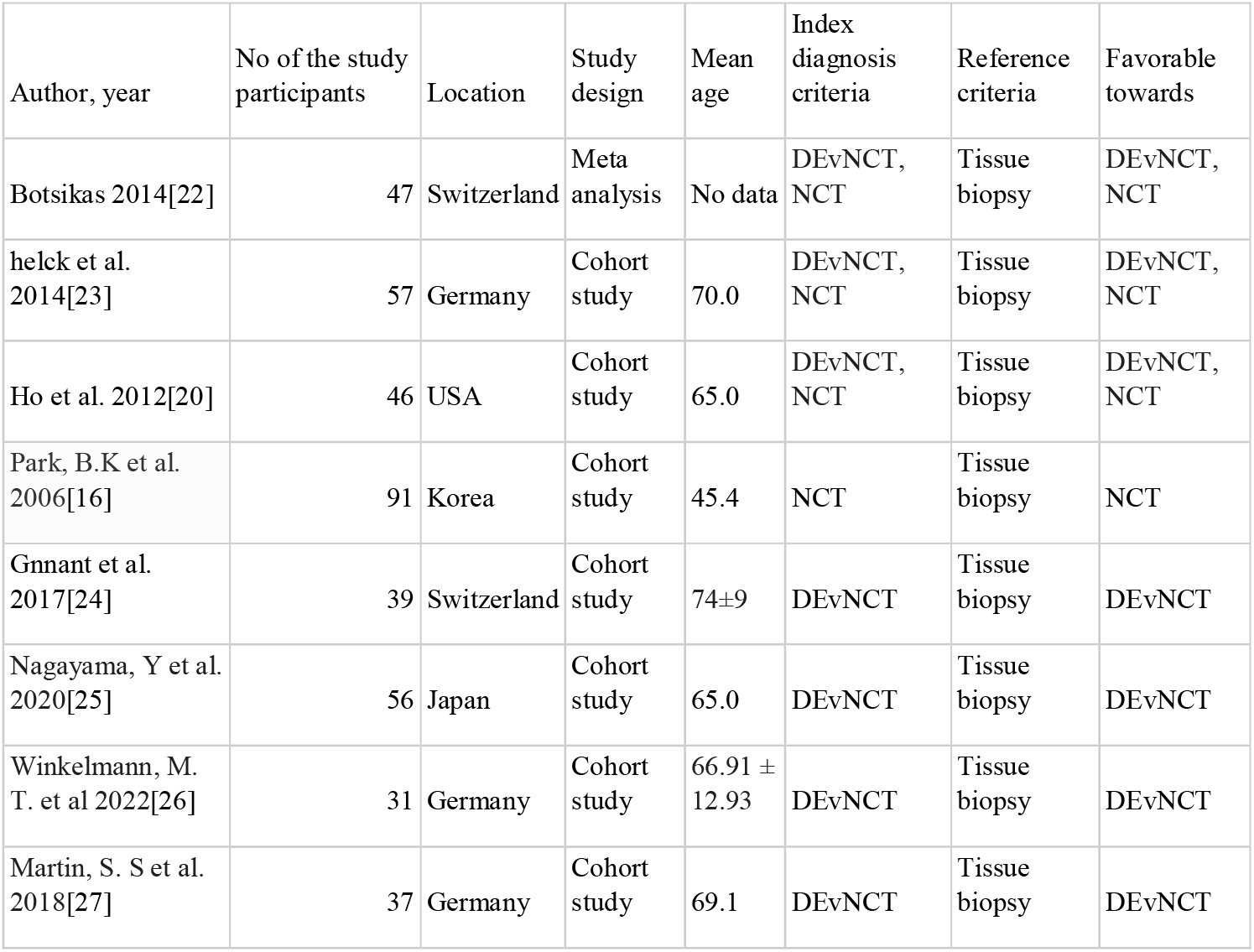
Table of Description of papers.

| Author, year | No of the study participants | Location | Study design | Mean age | Index diagnosis criteria | Reference criteria | Favorable towards |
| --- | --- | --- | --- | --- | --- | --- | --- |
| Botsikas 2014[22] | 47 | Switzerland | Meta analysis | No data | DEvNCT, NCT | Tissue biopsy | DEvNCT, NCT |
| helck et al. 2014[23] | 57 | Germany | Cohort study | 70.0 | DEvNCT, NCT | Tissue biopsy | DEvNCT, NCT |
| Ho et al. 2012[20] | 46 | USA | Cohort study | 65.0 | DEvNCT, NCT | Tissue biopsy | DEvNCT, NCT |
| Park, B.K et al. 2006[16] | 91 | Korea | Cohort study | 45.4 | NCT | Tissue biopsy | NCT |
| Gnnant et al. 2017[24] | 39 | Switzerland | Cohort study | 74±9 | DEvNCT | Tissue biopsy | DEvNCT |
| Nagayama, Y et al. 2020[25] | 56 | Japan | Cohort study | 65.0 | DEvNCT | Tissue biopsy | DEvNCT |
| Winkelmann, M. T. et al 2022[26] | 31 | Germany | Cohort study | 66.91 ± 12.93 | DEvNCT | Tissue biopsy | DEvNCT |
| Martin, S. S et al. 2018[27] | 37 | Germany | Cohort study | 69.1 | DEvNCT | Tissue biopsy | DEvNCT |

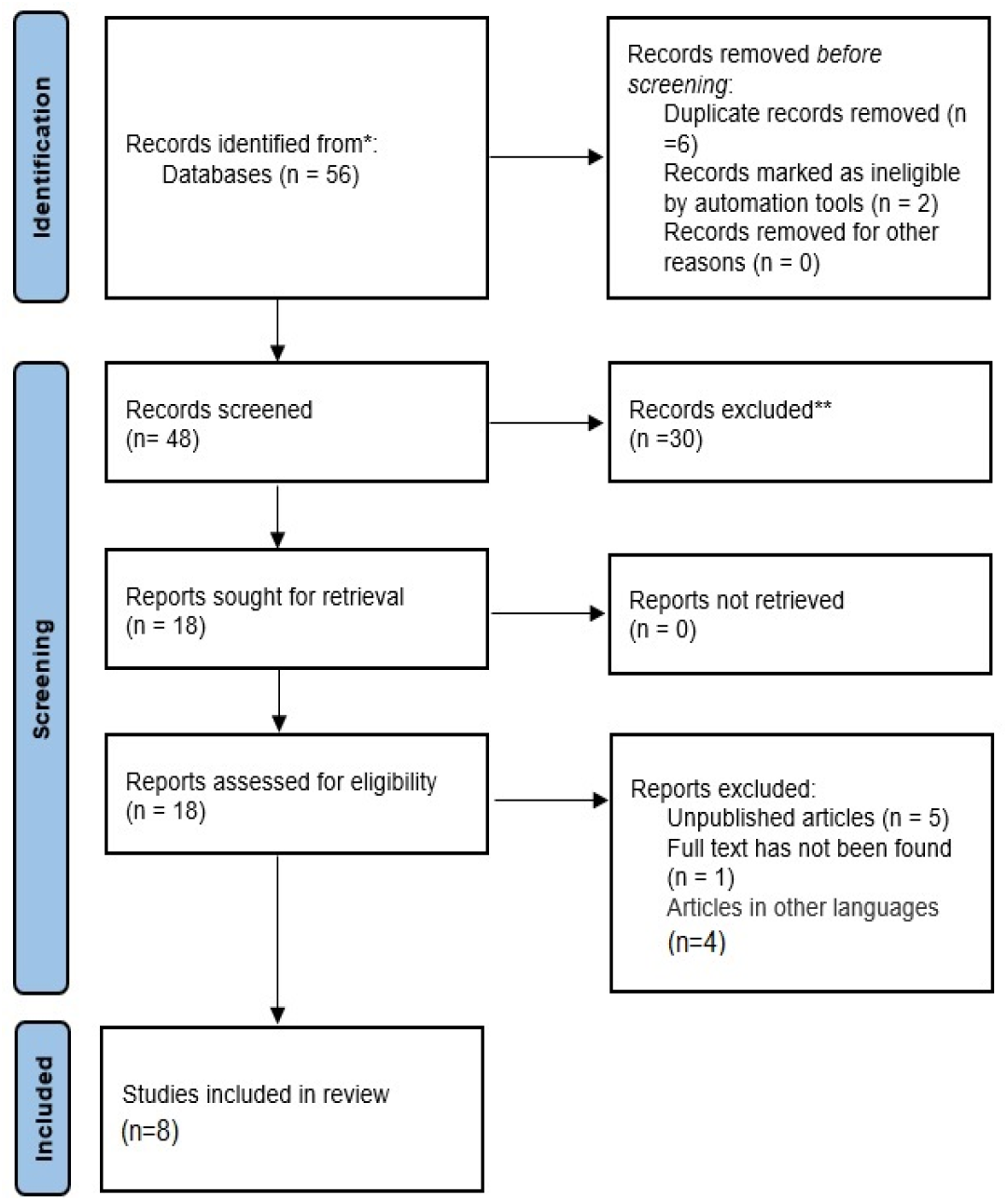
PRISMA Flow Chart (Figure 1)

**Figure 2:**
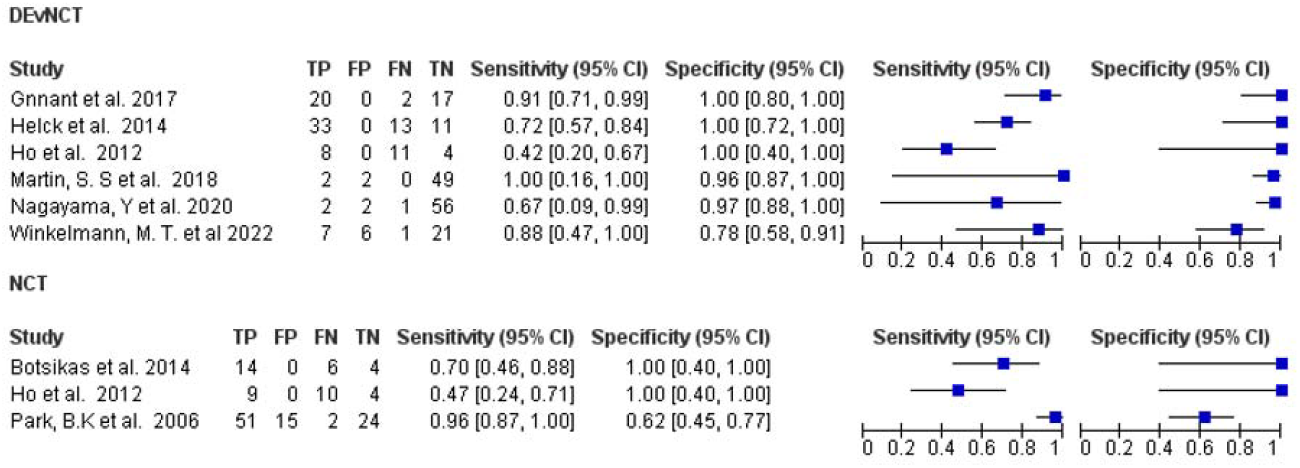
The forest chart summary for DEvNCT and NCT.

**Figure 3:**
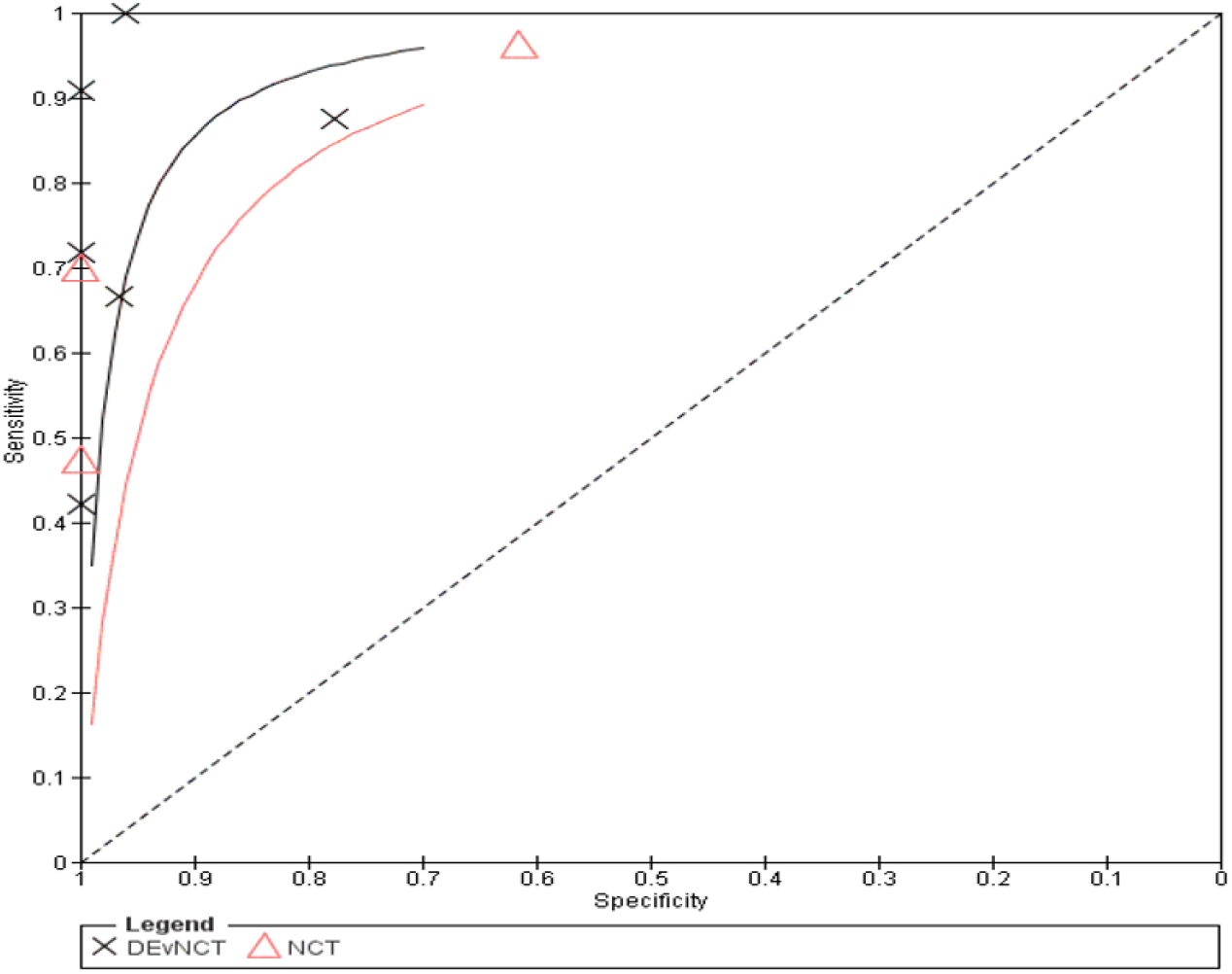
The SROC plot summary for DEvNCT and NCT.

The summary of the ROC curve is described in Figure 3. It shows that the area under the ROC (AUC) for DECT was 0.8302 and the overall diagnostic odds ratio (DOR) was 40.629. It also describes the Diagnostic Accuracy and the younden index. which are 0.858 and 0.66 respectively.

Figure 4 describes the summary of Fagan plots analysis for all the studies of DECT vs NCT, it shows a Prior probability of 37% (0.6); a Positive likelihood ratio of 12; a probability of post-test 88% (7.1); a Negative likelihood ratio of 0.30, and a probability of post-test 15% (0.2).

**Figure 4:**
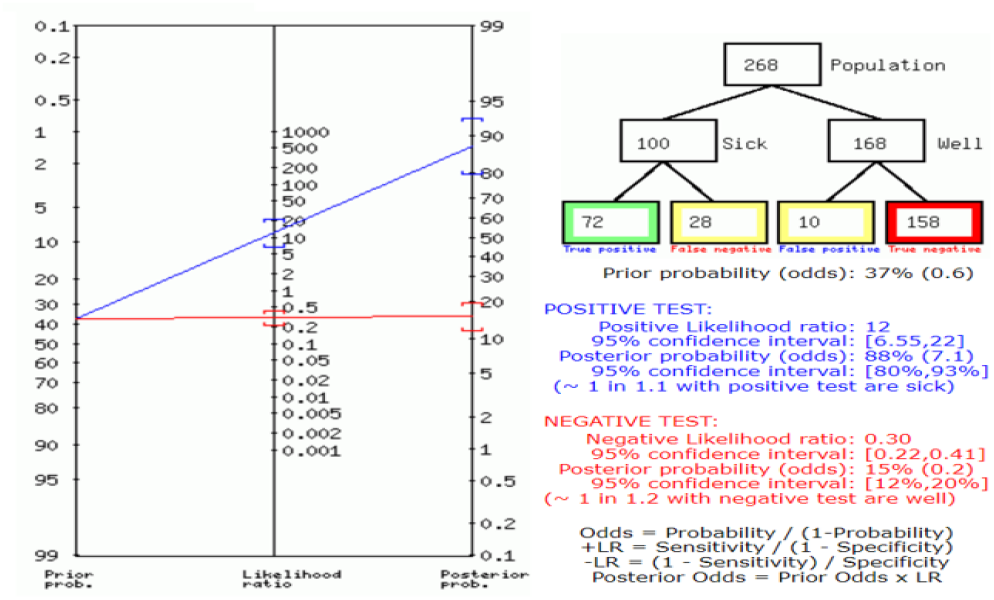
Fegan’s Analysis for DECT vs NCT.

### NCT vs Biopsy

Above, Table 1 Describes all the description of papers used for the NCTvBiopsy study. As the result described above, in the forest chart (Figure 2), the comparison of the sensitivity and specificity of different papers can be seen. The same is illustrated in the SROC curve (figure 3). A total of 3 RCTs with 139 patients were selected for the study. Here, 1 study showed sensitivity above 95% and 2 studies showed specificity above 95%. The value of True positive was 74, True Negative was 32, False negative was 18, and False Positive was 15. With a confidence interval of 95%, Sensitivity, specificity, and Positive Predictive values were calculated. A summary of this is available in Figure 2. The Sensitivity of the NCT is 0.804. with a CI of 95% in a range of (0.528 to 1.081); the mean being (0.277). The Specificity of the NCT is 0.681 with a CI of 95% in a range of (0.43 to 0.932) the mean being (0.251). The positive predictive value (PPV) for NCT is 0.831 with a CI of 95% in a range of (0.683 to 0.98) the mean being (0.148).

The summary of the ROC curve is described in Figure 3. It shows that the area under the ROC (AUC) for the NCT was 0.742599 and the overall diagnostic odds ratio (DOR) was 8.77. Also, The Diagnostic Accuracy and younden index are 0.763 and 0.485 respectively.

Figure 5 describes the summary of Fagan plots analysis for all the studies of NCT vs Biopsy, it shows a Prior probability of 66% (2.0); a Positive likelihood ratio of 2.52; a probability of post-test 83% (4.9); a Negative likelihood ratio of 0.29 and a probability of post-test 36% (0.6).

**Figure 5:**
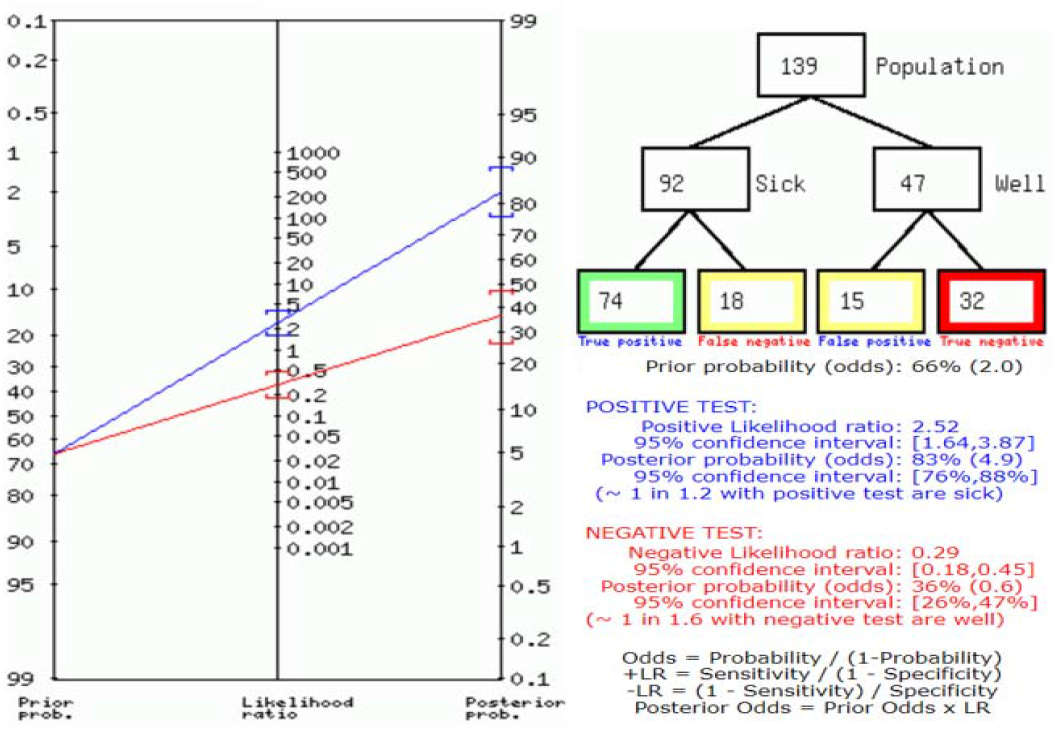
Fegan’s Analysis for NCT vs Biopsy.

### Bias Study

#### Publication Bias for DECT vs NCT and NCT vs Biopsy

The summary of publication bias for the DECT vs NCT and NCT vs Biopsy study is shown in (figure 6 and Figure 7). For the publication bias, In, patient selection, bias was low in 8 studies and unclear in 1. In the index test, it was low in 7, high in 1, and unclear in 1 study. While the reference standard was low in 4, high in 2, and unclear in 3. The flow and timing were low in 7, and unclear in 2. The applicability concerns in patient selection were low in 7 and high in 1 and unclear in 1. The Reference standard was low in 2, high in 3, and unclear in 4. The index test was high in 2 and low in 7.

**Figure 6:**
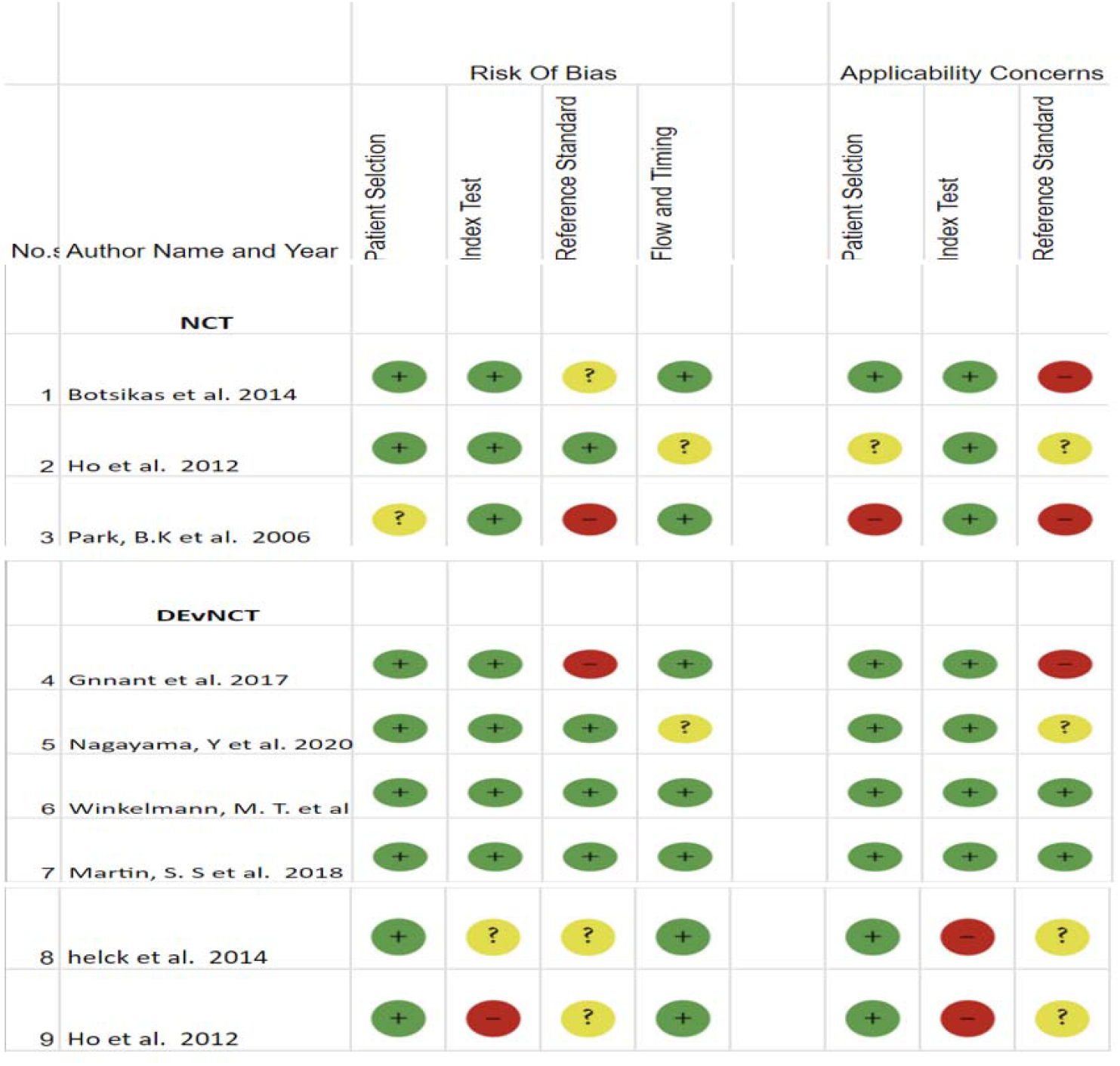
Bias Study.

**Figure 7:**
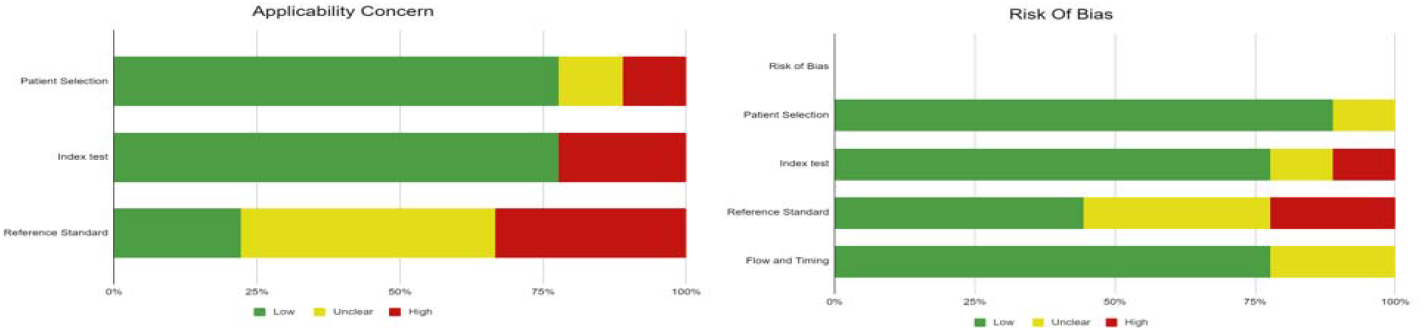
Summary of Applicability Concern and Risk of Bias.

## Discussion

Adrenal adenomas are generally benign neoplasms arising from the adrenal cortex. Non-secreting adrenal adenomas secrete low levels of hormones, and as a result are usually asymptomatic, typically being discovered incidentally on abdominal imaging.[28]

Combined enhanced and unenhanced computed tomography is the currently established method of choice for adrenal lesion description.[8] The protocol that is currently being followed for adrenal lesion characterization consists of an unenhanced acquisition wherein a lesion with a density lower than 10 HU is characterized as an adrenal adenoma. It is also pertinent to understand that when the adrenal lesion is incidentally found on an enhanced CT, with no unenhanced acquisition available at that time, the patient has to be recalled to perform a dedicated CT examination, leading to inconvenience as well as loss of time in the management of the lesion, if and when needed [29]Modern day studies have documented that there is no or statistically insignificant difference in adrenal mass attenuation between virtually unenhanced and enhanced images.

In this systematic review and meta-analysis, we found that vNCT images generated from dual-energy CT demonstrated comparable sensitivity to NCT for the diagnosis of adenomas. These findings are of potential importance because the diagnosis of adenomas using vNCT alone could prevent additional follow-up imaging studies and reduce cost and cumulative radiation dose (if NCT is pursued as the next imaging test) to the patient. Our analysis indicated that dual-energy vNCT had a sensitivity of 72% and a specificity of 94%, with a PPV of 0.878 from a total of 6 papers being analyzed, in comparison to a biopsy, which understandably is the best method to confirm a diagnosis of an adrenal adenoma, concluding it with evidence of histologic architecture. When 3 other papers were analyzed to study the impact of NCT in the diagnosis of incidental adrenal adenoma, it yielded a sensitivity of 80% with a specificity of 68.1%, along with a PPV of 0.831.

The analysis clearly indicates that while NCT is more sensitive by approximately 8%, DEvNCT functions with a specificity of 94%, a difference of more than 25% from that of NCT. This cements the understanding that while the difference in sensitivity may be circumvented or diluted for application, the difference in specificity is vastly tilted towards DEvNCT. This difference may in part be contributed to diversity in the ethnicity of subjects used, papers only being considered as per our inclusion and exclusion criteria, as well as the fact that the papers being considered relevant for NCT are studies performed until 2014, while those being considered for DEvNCT have been conducted in the more recent past, the impact of which is currently beyond the scope of this study. To aid our understanding of these modalities and their impact, Kim et al. found that the mean attenuation value of the lipid-rich adenomas on DEvNCT images (11.7 HU ± 9.5) was significantly greater than those on NCT images (0.7 HU ± 7.2) (p = 0.001). This is a difference that is both statistically significant and can also influence the characterization of an adrenal adenoma, as the cut-off of 10 HU established thus far can be easily overpassed.[30]

That being said, it must also be noted that dual-energy CT permits the differentiation of materials on the basis of their energy-related attenuation characteristics by using two energy settings at the same time and also provides the ability to generate virtual unenhanced data setpoints from contrast-enhanced DECT images. [31], [32]. Studies such as Gupta et al. and Shi et al. also proposed that DECT can be used to help differentiate some lipid-poor adrenal adenomas from metastatic lesions based on their attenuation values. [33], [34]

Based on these findings, it is safe to assert that DEvNCT promises to be a more vital and impactful modality, with its impact and accuracy the closest to reaching that of a biopsy beyond its impact through its invasive nature. This study may be limited in its analysis due to the relatively lesser number of papers being considered through its stringent inclusion criteria, as well the barrier created by considering studies written only in the English language, as well as the discovery of scientifically relevant facts and findings that were not considered or updated when the previous studies were formulated.

## Data Availability

All data produced in the present work are contained in the manuscript

